# How far Covid19 virus spread can be curbed by relaxing lockdown in different stages? -A study in Indian scenario

**DOI:** 10.1101/2020.06.11.20129023

**Authors:** Arindom Chakraborty, Kalyan Das

## Abstract

After the emergence of the first cases in Wuhan, China, the novel coronavirus (2019-nCoV) infection has rapidly spread out to other provinces, neighboring countries and finally has become a global terror. It is indeed a matter of serious concern to study the transmission dynamics of this virus. The potential and severity of an outbreak and providing critical information for identifying the type of disease interventions and intensity can be well understood by the unknown basic reproduction number. A stochastic model can be used to estimate this number with possible safeguard on uncertainties. It is essential to assess how the expensive, resource-intensive measures can contribute to the prevention and control of the 2019-nCoV infection and how long they should be maintained. A short-term forecast of incidences are often of high priority. The challenge is to forecast unseen “future” simulated data for three different scenarios at some time points. We estimate current levels of transmissibility, over variable time points under different levels of interventions and use that to forecast near-future incidence. The forecasted values of incidence can be used for determining the near future mortality also.

## Introduction

It is always a challenge to work with reliable statistical models, for projecting infectious disease spread, for many reasons. It is expected that the model should posses a few qualities like transparency, reproducibility, validity etc. Modelling the spread of novel coronavirus(2019-nCoV) is no exception. Not only the researchers and policy makers, the public is also very keen to know when the mayhem caused by that particular disease will stop. People are gradually accepting the fact that this virus will stay with us for some more time and some non-pharmaceutical interventions will be imposed. In this situation, critical decisions will rely on these statistical models.

Ever since the emergence of the novel coronavirus (2019-nCoV), comprehensive and stringent interventions have been attempted to implement in order to decelerate the spread of the disease. The effectiveness of interventions still remain ambiguous. In the absence of pre-pandemic vaccines and antiviral drugs for the outbreak of novel pathogen COVID19, a potentially attractive policy option is based on nonpharmaceutical interventions. This requires substantial investments in reactive interventions, with consequent implementation plans. Social distancing and lockdown are the immediate solutions to check the outbreak. But that results in serious repercussions on the economy specially due to the absence of economic activities. The direct impact of lockdown is huge as recent study shows that the lost income of marginal workers (casual labourers and workers involved in MNREGA, a scheme related to guaranteed rural employment) is more than 10% of GDP (Gross Domestic Product) and that of consumption expenditure is around 2.5% of GDP if complete lockdown is strictly adhered for 60 days. This necessitates the study on the impact of relaxation of lockdown in different states and as a whole in India. The lockdown level may be viewed by the policy makers in terms of relaxation of transport, mobility, mask (gloves) use, the closure of schools and universities, banning of mass gatherings and/or public events, social distancing and others.

There have been substantial literature on the statistical prediction of the spread and mortality due to COVID19, eversince the outbreak occurred in Wuhan, China. Linton et al.^1^ (2020) consider the estimation of incubation period for this corona virus infection. Kiesha Prem and his colleagues^2^ and Ferguson et al.^3^ (2020) observe the effect of nonpharmaceutical interventions for reducing the mortality and infected cases. Tidman (2020)^4^ considers a case study to see the effect of blanket testing on the new incidents of infection.

The different levels of lockdown play a major role in slowing down the attacking rate of the virus. The main focus is therefore to reduce the effective reproduction number, *R*_*t*_ of the infection, a fundamental epidemiological quantity representing the average number of infections, at time t, per infected case over the course of their infection. If *R*_*t*_ is maintained at less than 1, the incidence of new infections decreases, ultimately resulting in control of the epidemic. If *R*_*t*_ is greater than 1, then infections will increase (dependent on how much greater than 1 the reproduction number is) until the epidemic peaks and eventually declines due to acquisition of herd immunity. Our target here is to study the pattern of this rate in states over the days under different levels of lockdown.

The stochastic model we consider is important for taking into account the uncertainty in prediction. Note here that we stress upon the behaviour of the reproduction number under different levels of lockdown over days. A Bayesian model of the infection cycle to observe deaths, inferring plausible upper and lower bounds (Bayesian credible region) of the total populations infected, case detection probabilities and the reproduction number over time. MCMC technique has been used for analyzing the data.

## Material & Methods

In this investigation essentially we have scaled different levels of lockdown (in the sense of restrictions imposed).The scales of lockdown are categories of a variable denoted as follows:

- No restriction (free movement) represented by category 0
- partial restrictions represented by category 1
- complete restrictions (as implemented in hotspots) represented by category 1

To predict the time varying reproduction number (*R*_*t*_) under various levels of lockdown, the reproduction number at time t is taken as a scale multiple of the baseline reproduction number. The multiplicity factor is a constant function of the lockdown levels. Note that we insert four dummy variables for the above four lockdown levels. The number of infected cases at any day can be predicted using the reproduction number and the weighted average of previous days’ affected figures with the discretized serial interval distribution probability of secondary infection as weights (See Fraser (2007)^5^, Cori et al. (2013)^6^, Nouvellete (2018)^7^, Cauchemez et al. (2008)^8^. The mortality at any day can similarly be predicted using the case fatality ratio (CFR) and the weighted average of previously affected figures with chance of mortality as weights.

Mathematically speaking, there are three models working together. These are models for infections, deaths and average reproduction rate. Let *I*_*s,t*_,*D*_*s,t*_ and *R*_*s,t*_ be, at time t, number of new infections, number of new deaths and average reproduction number for the state s. As explained above, we then express the infection on t th day as

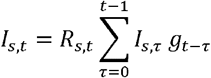

We discretize the serial interval as:

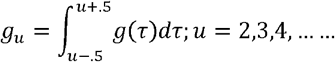

with

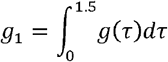

where from the past experience, g(t) (serial interval distribution) is assumed to be a gamma distribution with mean 6.5 (average time from onset in a primary infection to onset in a secondary infection) and a relatively small coefficient of variation0.62.

For mortality, the observed number of deaths may be assumed to follow a Negative Binomial law where the expected deaths are assumed to be the weighted average of the daily infection, weights being a mixture of two gamma distributions that accounts for the incubation period and time between onset of symptoms and death.

In case of *R*_*s,t*_, we use levels of lockdown as covariates. Let *R*_*s*,0_ be the baseline reproduction number for s-th state. Then *R*_*s,t*_ is modelled as

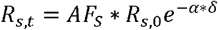

where *AF*_*s*_ is the adjustment factor considered for state s based on its population, *δ* indicates the level of lockdown 0, 1 or 2.

## Results

For each of states and India as a whole, predictions on number of daily infections, deaths, reproduction number upto 15th May, 2020 have been made. We have used four levels of lockdowns as ordinal covariates. For all the states we found, as expected, that continuing lockdown has significant role in controlling average reproduction number *R*_*t*_. We have also found the baseline reproduction number *R*_0_ for each state (Figure 1a). It is to be noted that while the country-wise *R*_0_ is estimated as 3.4036, for states like Gujarat *R*_0_ = 4:4672 and Rajasthan *R*_0_ = 3:5097 showed higher base line reproduction number. This indicates that these states may get worse if interventions are not taken seriously. In figure 1b, the recent *R*_*t*_ values are given. These values are for 31^st^ May, 2020. It is easy to conclude that this lockdown has played very important role in reduction of values for all the states. For Gujarat, the present *R*_*t*_ value has reduced to 1.3043 from 3.5680. Rajasthan has also showed improvement over time by reducing the same parameter value from 3.1730 to 1.1624. Similarly, for Delhi also we observed some improvement as the *R*_*t*_ value reduced to 1.4509 which was 3.9793 earlier. It was highest in the country. For Punjab, *R*_*t*_ started at 2.6379 which eventually reduced to <1 (0.9668). Due to high value of *R*_0_ (3.7146) for Tamil Nadu, lots of infections have been recorded. Current *R*_*t*_ value, as a result of prolonged lockdown, is calculated as 1.3607. Combining all states/union territories for India, the present *R*_*t*_ value is 1.2772 (as on 31^st^ May, 2020) which was 3.4872 on 14^th^ March, 2020.

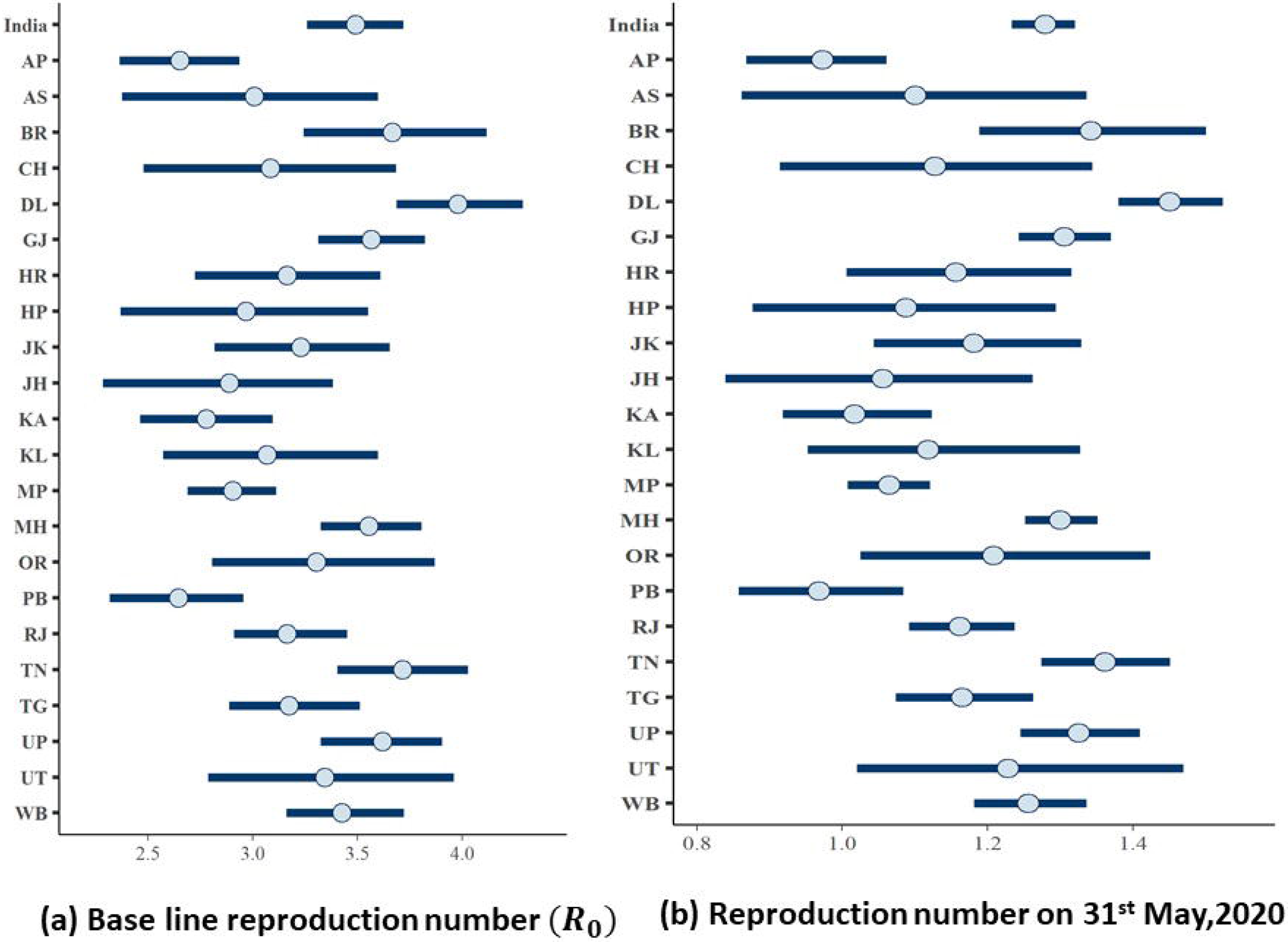

A lot of debates are going on about the extension of lockdown. Undoubtedly, this lockdown has long term implications in Indian economy. On the other side, it has already been shown that this lockdown has reduced the spread to some extent by controlling the value of average reproduction number *R*_*t*_. Here we have made an attempt to compare scenarios under different levels of lockdown starting from free movement to continuation of lockdown. We considered another possibility of partial lockdown keeping in mind the adverse effect of lockdown on economy.

For India, it may be noted that in all three situations, an increase trend is observed (Figure 2a-2c) in the number of daily new infections. However, in the prediction part, we have noticed steeper inclination of the curve after 4th May, 2020 if lockdown is withdrawn. This rate reduces with the increase in restrictions. Figure 2c shows flatter trend comparing to other situations. Same features have been observed in case of other states like Delhi (Figure 3a-3c) and Maharashtra (Figure 4a-4c). We have seen similar trend in all situations for other states too. It has also been reported that Kerala has achieved an incredible control over this COVID-19 spread. We have also tried to predict the scenario for Kerala (Figure 5a-5d). Our prediction shows that even under worst condition (no extension of lockdown after 31^st^ May, 2020), the daily number of newly infected is much less compared to other states and *R*_*t*_ is found to be 0.4921.

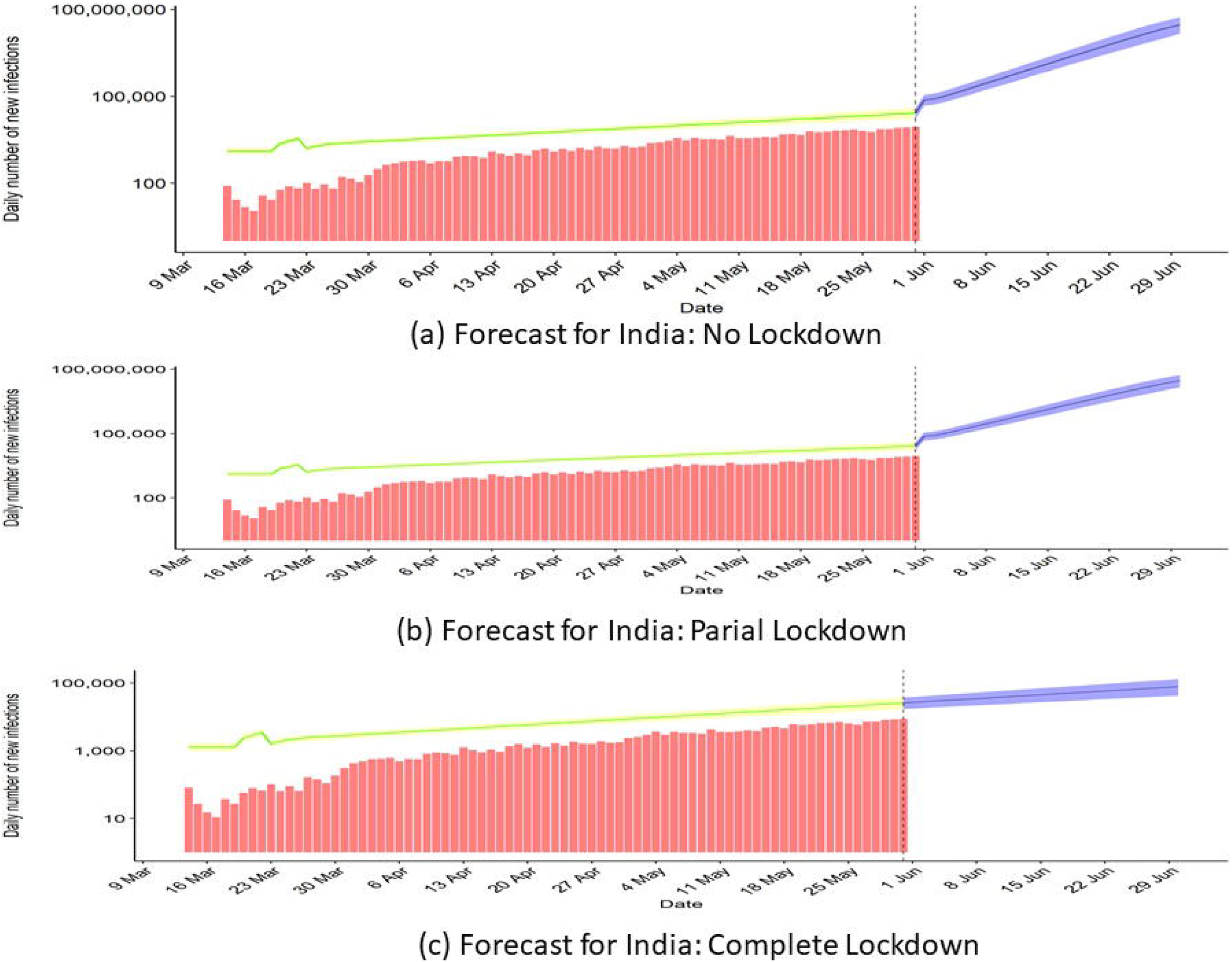

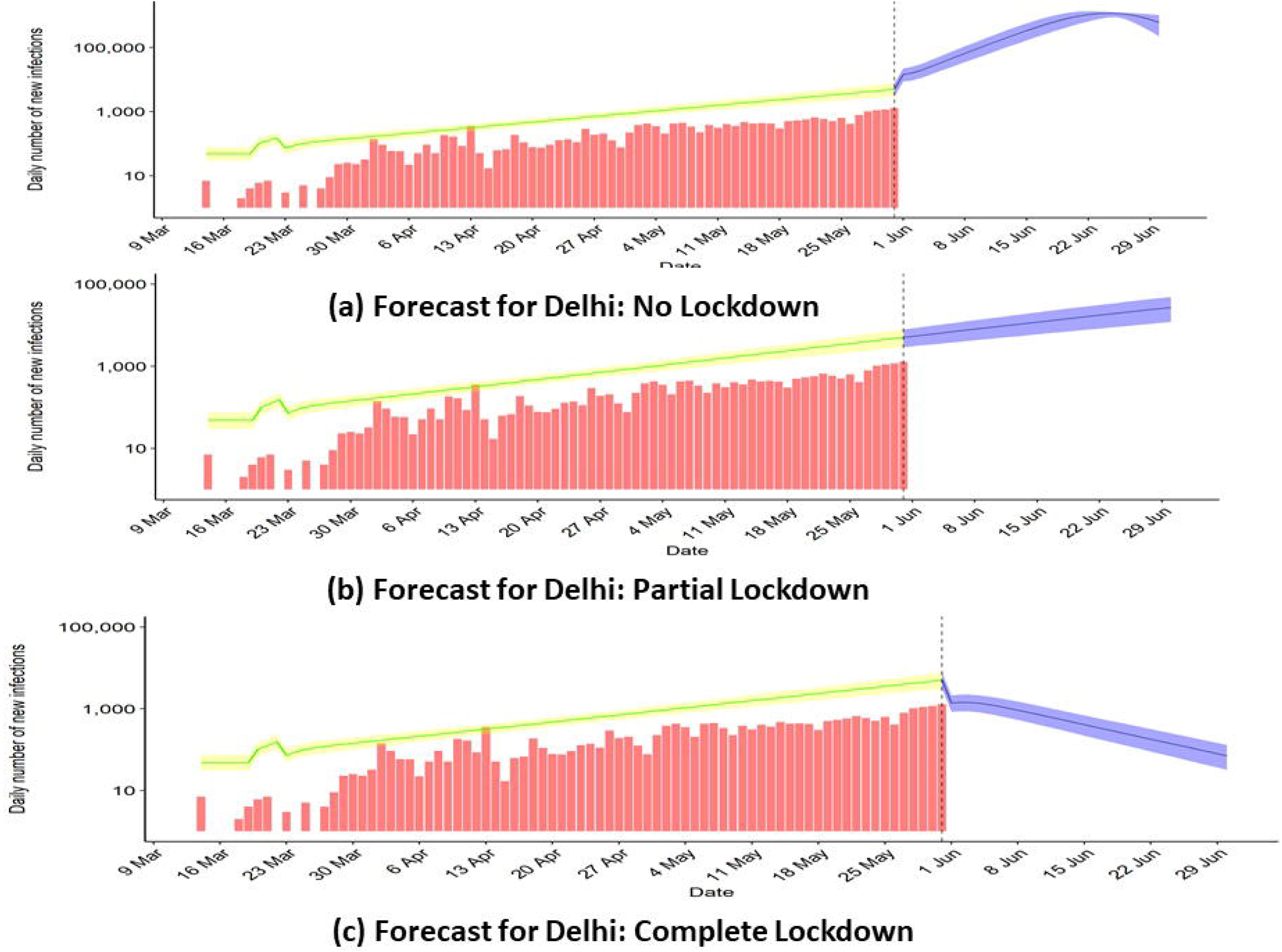

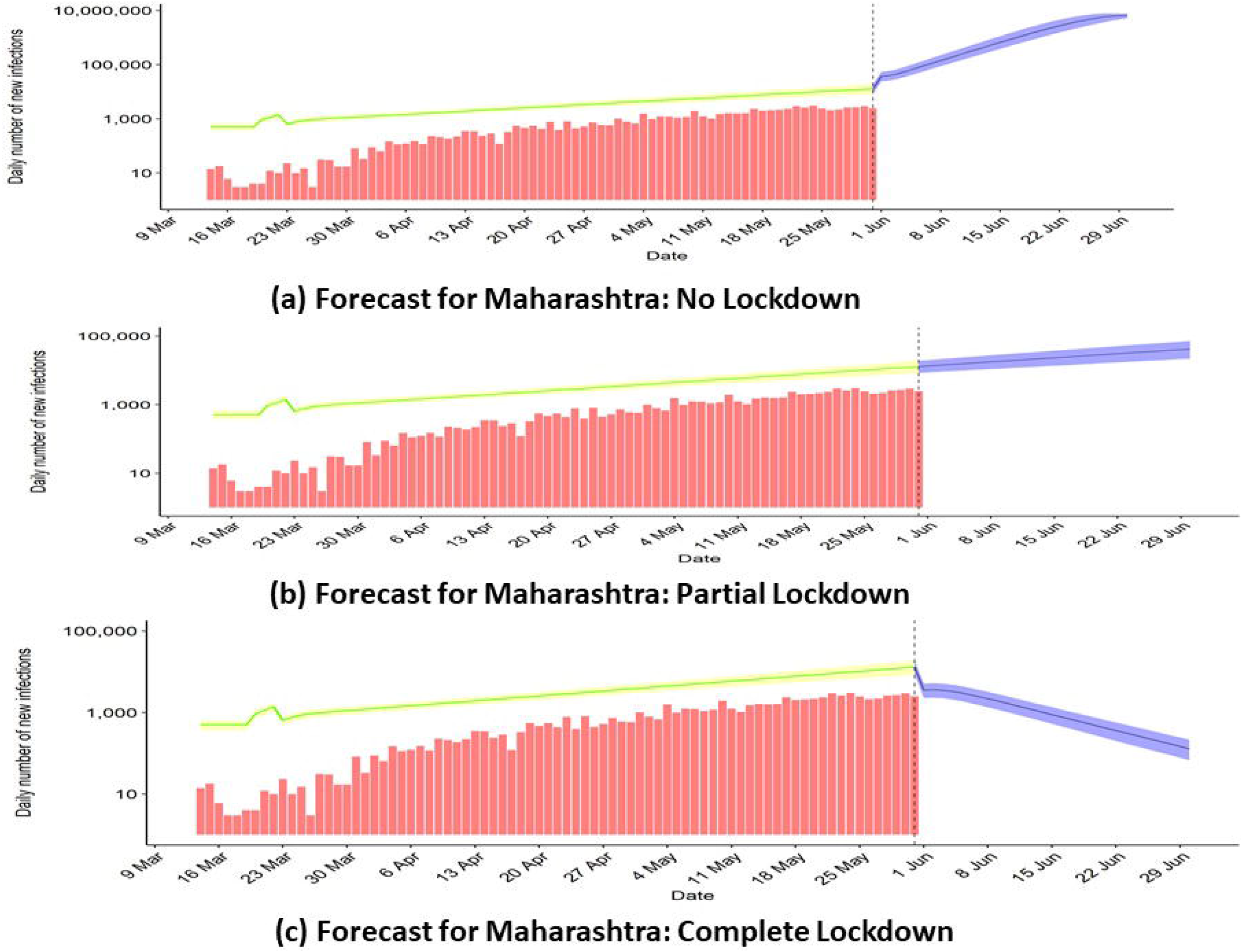

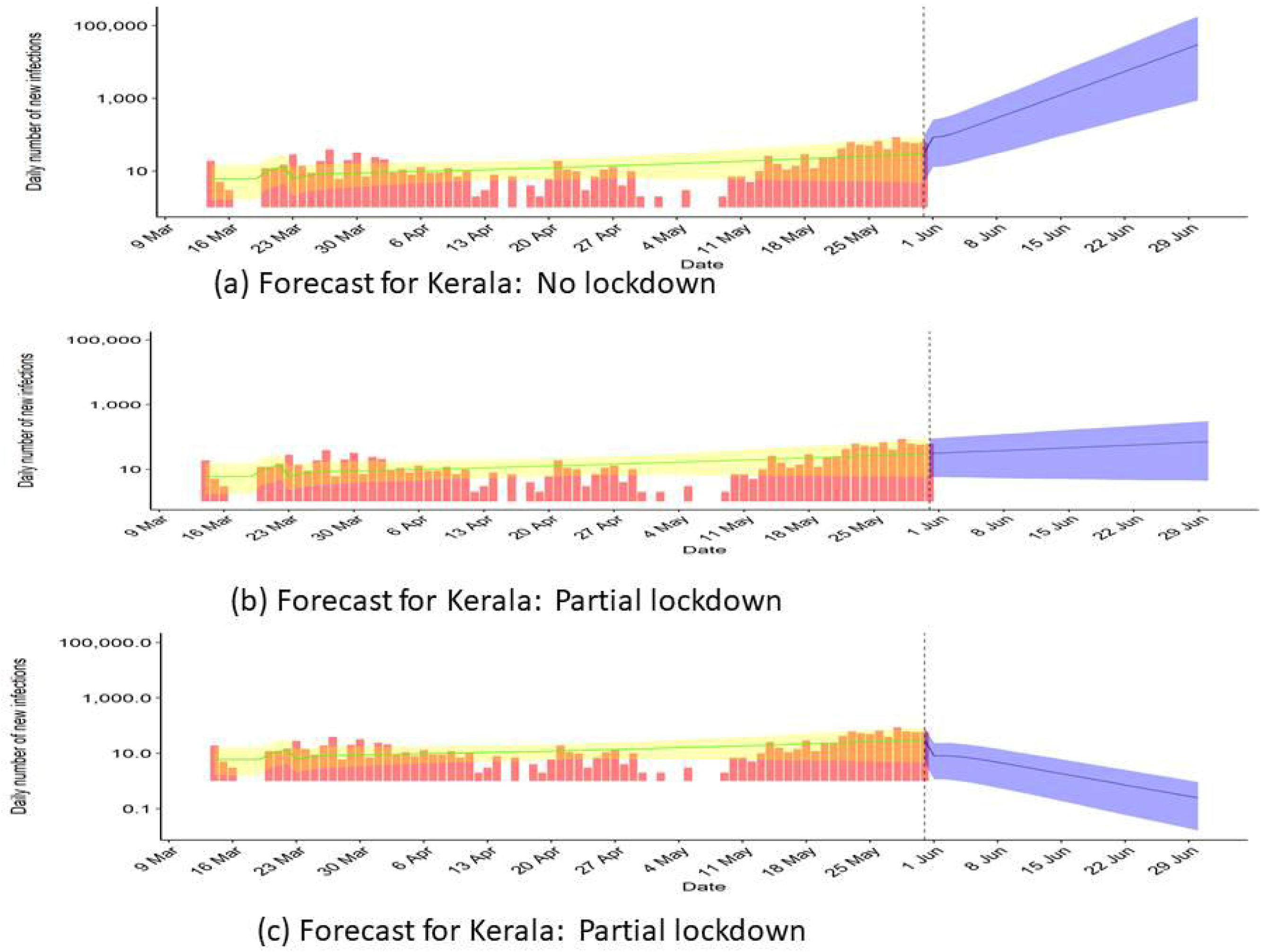

In general the mortality for COVID-19 is small compared to other diseases like SARS, MAERS. From Wuhan data, it is found that it has higher fatality among aged and persons with diseases like COPD, CVD, diabetes etc. Using the same model, we have also tried to predict the number daily deaths under the above mentioned set of lockdown levels. For India (Figure 6a-6c) we found that the daily death may cross thousand mark during the middle of June, if lockdown is lifted after 31^st^ May, 2020 (Figure 6a). If the lockdown is implemented completely, we may achieve the target of <100 daily death within third week of June (Figure 6c) and it will reduce after that. The states like Delhi, Maharashtra, Gujarat and West Bengal have also shown similar type of predictions. The gradual increment of restriction is helping in reducing the number of daily deaths. For Gujarat, however number of deaths predicted is a bit on the higher side in comparison. This may be attributed to the fact that average reproduction rate for Gujarat, till date, is on the higher side in the country resulting in more infected people.

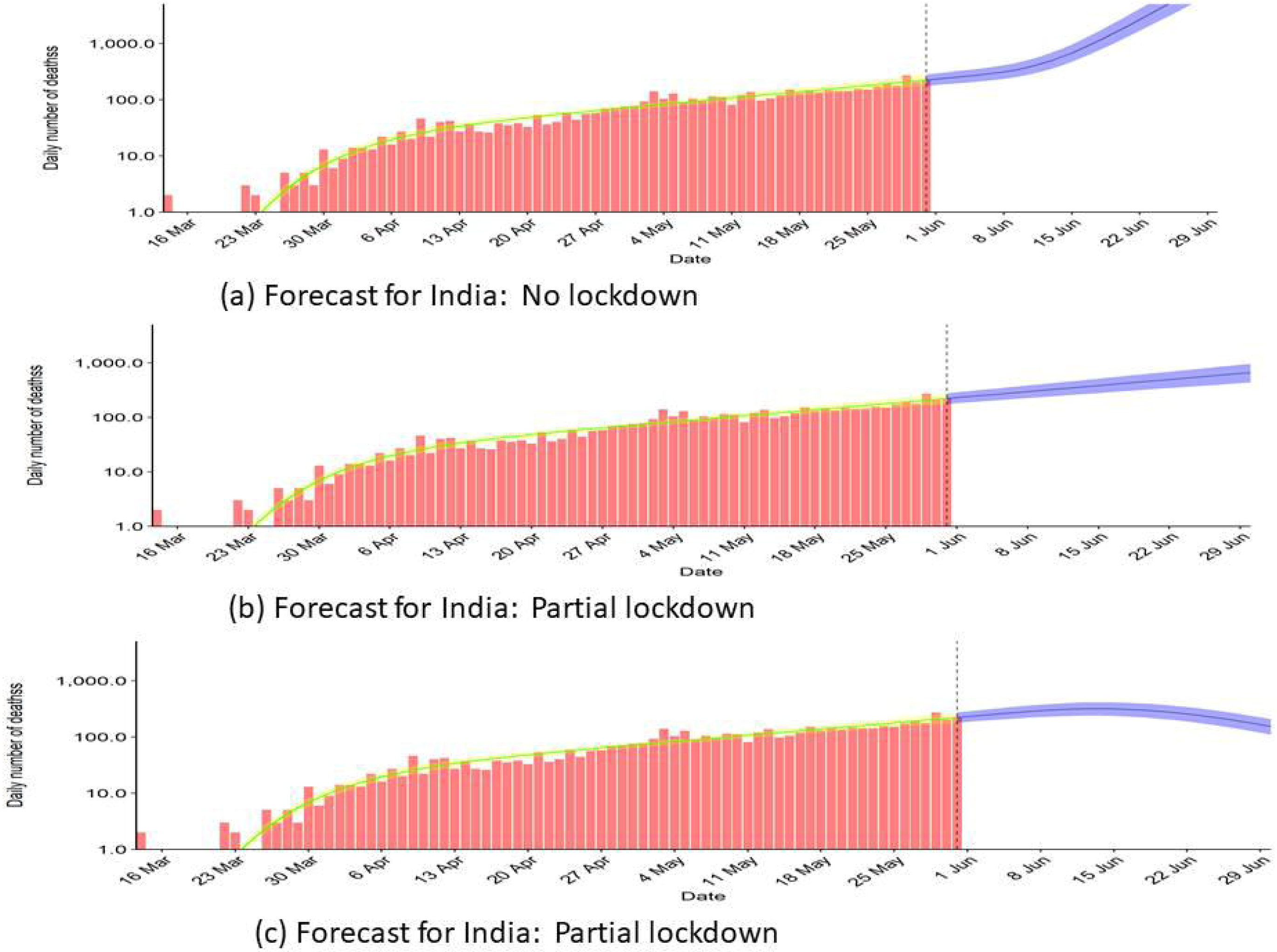

## Discussion

Over the last few months, there has been an explosion in the development of mathematical models for outbreaks of COVID19. In fact, the challenge is to make sure that these models represent reality. We intend to produce accurate trends while correctly accounting for uncertainties surrounding both the data and the transmission dynamics of the disease. One of the essential mathematical tools for understanding disease dynamics is to obtain the basic reproduction number. In the present study we consider a Bayesian hierarchical model to calculate that number. This number plays the major role in calculating the number of new incidents and the number of mortality in each day.

Availability of sufficient information particularly at state levels is a primary necessity for accurate prediction. As more data emerge all relevant parameters that influence transmission of this novel corona virus can be estimated steadily and hence models can more accurately predict the success or failure of different strategies to control the epidemic and limit mortality. The purpose may be to see the effect of all the items (like Road, rail and Air transport, proportion of mask (gloves) users, the closure of schools and universities, banning of mass gatherings and/or public events) that contribute to different levels of Lockdown, had there been data available at micro level on each specific item. To prevent health-care system overload in an already mounting epidemic, alternative strategy to distancing lockdown measures (after the relaxation of lockdown conditions) should be to incorporate testing, contract tracing, and localised Quarantine of suspected cases. Modelling such a strategy (like what proportion of the residents to be tested and how regularly testing could be done given asymptomatic and presymptomatic transmission) would be extremely useful to guide for controlling the pandemic when such measures could be lifted. Currently we are working on that and we plan to communicate our observation as soon as possible. Given that disease dynamics are greatly influenced by human behaviour, it seems in fact quite natural to have a human component in infectious disease modelling.

Imposing partial lockdown is a policy decision and may be subjective. The administrations, local as well states, have already started to relax the lockdown to revive the economy. Increasing mobility may result in more infections but definitely it will help the economy. So a trade-off may be done using the concept of partial lockdown.

Identifying the most effective control measures to reduce transmission in the community is indeed a challenging task. The characteristics of cases should continue to be monitored to find any changes in epidemiology — for example, increases in infections among persons in younger age groups or health care workers. Future studies could include forecasts of the epidemic dynamics and special studies of person-to-person transmission in households or other locations would be valuable. We hope that our investigation would help to provide a better insight on the spread of the virus. We believe the work would lead to some new avenues for future research.

## Data Availability

Data is available in public domain.

https://api.covid19india.org/

## Data

For this work we have used the data available in https://github.com/covid19india/api

## Additional information

### Competing interests

The authors declare no conflict of interests.

## Notes

### Competing Interest Statement

The authors have declared no competing interest.

### Funding Statement

No founding was available for this research

## References

1. Linton, N. M. et al. Incubation period and other epidemiological characteristics of 2019 novel coronavirus infections with right truncation: A statistical analysis of publicly available case data. J Clin Med. 9(2), 538, DOI: doi:10.3390/jcm9020538 (Feb 17, 2020).

2. Prem, K. et al. The effect of control strategies to reduce social mixing on outcomes of the covid-19 epidemic in wuhan, China: a modelling study. Lancet Public Heal. S2468-2667(20), 30073–6, DOI: doi:10.1016/S2468-2667(20)30073-6 (2020, Mar 2).

3. Ferguson, N. M. et al. Impact of non-pharmaceutical interventions (npis) to reduce covid19 mortality and healthcare demand. https://mcacs.org/multimedia/files/COVID19.pdf (2020).

4. Tidman, Z. Coronavirus: Italian village reports no new infections for days after blanket testing. https://www.independent.co.uk/news/world/europe/coronavirus-vo-euganeo-blanket-testing-venetoluca-zaia-a9411201.html (mMarch 19, 2020).

5. Fraser, C. Estimating individual and household reproduction numbers in an emerging epidemic. PLoS One 2(8), e758. DOI:doi:10.1371/journal.pone.0000758 (2007 Aug 22).

6. Cori, A., Ferguson, N. M., Fraser, C. & Cauchemez, S. A new framework and software to estimate time-varying reproduction numbers during epidemics. Am. J. Epidemiol. 178, 1505–1512, DOI: doi:10.1371/journal.pone.0000758 (2013).

7. Nouvellet, P. et al. A simple approach to measure transmissibility and forecast incidence. Epidemics 22, 29–35, DOI:doi:10.1016/j.epidem.2017.02.012 (2018).

8. Cauchemez, S., Valleron, A. J., Boëlle, P. Y., Flahault, A. & Ferguson, N. M. Estimating the impact of school closure on influenza transmission from sentinel data. Nature 452, 750–754, DOI: doi:10.1038/nature06732 (2008).

